# The landscape of coagulation cascade in pulmonary arterial hypertension

**DOI:** 10.1101/2024.03.18.24304511

**Authors:** Genfa Xiao, Guili Lian, Xiaoli Liu, Liangdi Xie

## Abstract

**Background:** Both the thrombotic and bleeding events were frequently complicated in pulmonary arterial hypertension (PAH), making recommendations regarding anticoagulation difficult.

**Methods:** The histopathological examination was performed on the lungs of two PAH models and global coagulation cascade alteration in lung tissue and peripheral venous blood was assessed by RNA sequencing and immunoassay, respectively. The clinical data and plasma samples were collected from PAH patients and controls and plasma coagulation cascade in subject was quantified by both immunoassay and proteomic approach.

**Results:** RNA sequencing analysis of lung tissues in two PAH models showed the reduced anticoagulants and intrinsic clotting factors and increased tissue factor (TF) expression. The immunoassay assessment of coagulation cascade in PAH models revealed increased TF expression, reduced intrinsic and common clotting factors and decreased anticoagulants antithrombin (AT)-Ⅲ in the peripheral circulation. Additionally, clinical evaluation of hemostatic parameters in PAH patients demonstrated hemostatic deficiency and lower AT-Ⅲ activity. Consistently, proteomic and immunoassay analysis of coagulation cascade in PAH patients revealed the increased extrinsic clotting factors, impairment of intrinsic and common pathways and impairment of AT-Ⅲ anticoagulant pathway.

**Conclusions:** The activation of extrinsic pathway and impairment of heparin/AT-Ⅲ anticoagulant system are conserved mechanisms for thrombosis across rat and human PAH, restoring its function with heparin supplementation may be a better option for future anticoagulant therapy.

**Clinical Perspective:** **What Is New?**

- The activation of extrinsic pathway and impairment of heparin/AT-Ⅲ system were responsible for thrombosis in pulmonary arterial hypertension.
- Impairment of intrinsic and common pathways, due to reduced plasma levels of clotting factors F10, F11, F2 and vWF, contributed to hemorrhagic complications, such as hemoptysis in PAH patients.

**What Are the Clinical Implications?**

- Increased risk of bleeding for anticoagulation with warfarin and novel oral anticoagulants may be attributed to further impairment of intrinsic and common pathways in PAH.
- Restoring function of heparin/ AT-Ⅲ anticoagulant system with heparin supplementation may be a better option for future anticoagulant therapy.

## Introduction

Pulmonary arterial hypertension (PAH) is a cardiopulmonary vascular disease characterized by elevated pulmonary arterial pressure and pulmonary vascular remodeling, leading to right ventricular failure and premature death. There was a high prevalence of thrombotic lesion in PAH ― pathologic studies showed thromboembolism and *in situ* thrombosis of the pulmonary vessels were the prominent findings with a frequency of 30–60%^1–4^. Congenital heart disease (CHD) is the most common contributing etiology of PAH in China. Patients with cyanotic CHD, have a high incidence of cerebral and pulmonary thrombosis of around 30%–40%^5^. Thromboembolic complications substantially contribute to the increased morbidity and mortality in PAH patients^6^.

Notably, current data on the thrombus formation in PAH were confusing. On one hand, hypercoagulable state has been suggested in PAH and associated with the increased risk of developing thrombus^7, 8^, on the other hand the lack of evidence for increased thrombus generation were reported^9–11^. Moreover, hypocoagulable state and multiple haemostatic defects have been demonstrated in patients with PAH^12–14^. Converse to thrombosis, the hemorrhagic events, such as hemoptysis were frequently complicated in patients with PAH, particularly to PAH associated with CHD and connective tissue disease^15–17^. In light of the high prevalence of thrombosis in the distal pulmonary arterioles, anticoagulant therapy would be beneficial for PAH patients. However, the potential anticoagulant benefits with warfarin have been impaired by bleeding events in the large-scale clinical trials^18^. These bleeding complications make recommendations regarding anticoagulation therapy difficult. Thereby, no consensus has been made for the optimal anticoagulant therapy and individual decision-making has been recommended by the 2022 ESC/ERS clinical guideline^19^.

The coagulation cascade is the process by which the body forms blood clots to prevent excess blood loss and culminates in the formation of thrombin, an enzyme responsible for the conversion of soluble fibrinogen to the insoluble fibrin clot. The formation of fibrin clot is the consequence of activation of extrinsic, intrinsic and common pathway, which is antagonized by three major natural anticoagulant mechanisms: tissue factor pathway inhibitor (TFPI) pathway, the protein C anticoagulant pathway and the heparin/antithrombin (AT) -*Ⅲ* pathway. In addition, fibrinolytic system also plays an important role in preventing clot propagation and removing already existed thrombus, inhibition of which promotes thrombus formation. Previous studies involved in thrombosis in PAH were rare and the sample sizes were relatively small. Only limited abnormalities in coagulation cascade were reported, including increase in plasminogen activator inhibitor 1 (PAI1)^9^ and tissue-type plasminogen activator (tPA) and plasmin-anti-plasmin^10^, decrease in soluble thrombomodulin (TM)^20^, and abnormality in clotting factors, including von Willebrand factor (vWF), tissue factor (TF) and fibrinogen^9, 14, 21, 22^. The mechanisms underlying thromboembolic and hemorrhagic complications in patients with PAH were largely unknown.

No study to date has thoroughly explored the global coagulation cascade alteration in PAH. The complexity of coagulation cascade, comprised of coagulation system, natural anticoagulant system and fibrinolytic system and over 40 components, may explain absence of such studies. In the present study, we performed an analysis of whole coagulation cascade in both lung tissues and peripheral circulation, aiming to identify the mechanisms underlying *in situ* thrombosis of pulmonary vessels and thromboembolic complications in PAH and provide therapeutic strategy for future anticoagulation.

## Methods

### Data Availability

The RNA sequencing datasets presented in this study can be found in the gene expression omnibus (GEO) repository, https://www.ncbi.nlm.nih.gov/geo/. The accession number(s) were GSE149713, GSE149899 and GSE8078, respectively. The proteomic and transcriptomic data generated in this study will be deposited to public repositories at the time of publication. Requests for proteomic and transcriptomic data and individual clinical data can be made via email to the corresponding authors. Other data generated or analyzed during this study are included in this manuscript and its supplementary information files.

### Animal and treatment

Adult SD rats were purchased from Shanghai SLACCAS Laboratory Animal Co., Ltd. The approval number is SCXK (hu) 2022-0004. The rats were raised with food and water ad libitum in the animal room. The monocrotaline (MCT)-induced PH model was established by a single intraperitoneal injection of 40 mg/kg MCT (Sigma-Aldrich, CA, USA). For Su5416+hypoxia (SuHx) -induced pulmonary hypertension (PH) model, rats were received a single subcutaneous injection of 20 mg/kg Su5416 (Glpbio, CA, USA) and then exposed to a 10% oxygen hypoxic chamber for 3 weeks before being returned to normoxia (21% O_2_), and for another 2 weeks in normoxia. The 10% O_2_ concentration was regulated by Oxygen Controller (Yuyang S1007, Shanghai Yuyan Instruments Co.,Ltd., China). The concentration of CO_2_ was maintained below 1000ppm using medical soda lime CO_2_ absorbent (Shanghai Nahui Desiccant Factory, China). Relative humidity within the chamber was maintained 40-60% using anhydrous CaCl_2_ (Xilong Chemical Co., Ltd., China), and the NH_3_ level was controlled using boric acid (Xilong Chemical Co., Ltd., China). Control rats were injected with vehicle and maintained at room air for 5 weeks. Control and MCT-treated rats were sacrificed at the end of week 1, 2, 3 and 4 after MCT treatment, while the hypoxic rats were sacrificed at the end of week 5 after injection of Su5416. After the hemodynamic evaluation, the venous blood samples were collected in EDTA vacuum tubes, and then plasma samples were obtained by centrifugation at 3000 g for 10 minutes at 4°C and stored at −80°C. All efforts were made to diminish suffering by using sodium pentobarbital anesthesia. The animal studies and procedures were approved by the Laboratory Animal Welfare and Ethics Committee of Fujian Medical University (Approval No. IACUC FJMU 2022-0842).

### Histopathologic examination

The histopathological examination was performed on sacrificed SD rats after blood collection. The chests of rats were opened and the lower lobe of right lung was dissected for hematoxylin and eosin (HE) staining. The lungs were fixed with 10% neutral buffered formalin for 24h, then washed with tap water. After dehydration by different concentration of alcohol and immersed in xylene, the lungs tissues were embedded in paraffin and sectioned at 3 μm thickness. Tissue sections were deparaffinized in xylene solution, rehydrated through 100 to 50% gradient alcohol, and rinsed in running tap water for 10 min. Then the specimens were stained with hematoxylin and eosin, and sealed with neutral balsam mounting medium. The samples were observed by microscope (Nikon, Japan).

### RNA extraction, cDNA library preparation and RNA sequencing

The left lung tissues of control and hypoxic rats were isolated, frozen in liquid nitrogen and stored at −80°C after sacrifice. RNA extraction, quality assessment, cDNA library preparation and RNA sequencing were described in our previous study^23^. Total RNA with high quality was used for cDNA library preparation. cDNA Library preparation and RNA sequencing were performed on an Illumina HiSeq 2000 platform by Biomarker Biotechnology (Beijing, China) Co., Ltd. The raw sequencing data were processed by a series of steps: (a) the removal of low-quality reads and adapter sequences by Skewer software; (b) the quality control by FastQC software; (c) the alignment of clean reads to rat reference genome by HISAT2 software; (d) the assembling of transcripts by the StringTie software; (e) The determination of transcript abundance by StringTie software through calculation of fragments per kilobase of transcript per million fragments mapped.

### Data preparation and bioinformatics analysis

The raw sequencing data of MCT-induced PAH were derived from our previous study and deposited in Gene Expression Omnibus (GEO) with an accession number GSE149713. The detailed methods regarding animal grouping and treatment, lung tissue and RNA isolation, cDNA library preparation and RNA sequencing were described in our previous study^23^. Additionally, two independent datasets with accession number GSE149899 and GSE8078 were downloaded from the gene expression omnibus (GEO). Morpheus, a versatile software, was used for hierarchical clustering analysis and heatmap creation (https://software.broadinstitute.org/morpheus/).The genes annotated in coagulation cascade were retrieved from KEGG pathway database (https://www.kegg.jp/entry/pathway+rno04610).

### Patient enrollment and blood sample collection

We recruited the treatment-naive patients with CHD and healthy volunteers from January 1, 2020 to January 1, 2024. In total, 91 consecutive patients with CHD and 35 healthy volunteers were recruited in this study. Pulmonary arterial hypertension is defined by a mean pulmonary arterial pressure (mPAP) >20 mmHg at rest^19^. Accordingly, 54 CHD patients were complicated with PAH by right heart catheterization. In addition to CHD, the other contributing etiologies of PAH were not included. The exclusion criteria included acute infection, concurrent coagulopathy, hematologic disorder, receiving targeted therapy and anticoagulant therapy. Healthy volunteers were enrolled from staff and blood donors. The healthy volunteers admitted neither history of cardiac or pulmonary diseases nor drug therapies. Peripheral venous blood was collected at admission. Plasma samples were obtained by centrifugation at 2000 rpm for 10 minutes at 4°C and stored at −80°C within three hours from blood collection. This study protocol was approved by the ethics committee of First Affiliated Hospital of Gannan Medical University and written informed consent was obtained from each participant (Approval Number LLSC-2023No.235).

### Clinical data collection

We reviewed the medical records of CHD patients who underwent right heart catheterization to assess cardiac and pulmonary haemodynamics. The details about haemodynamics, including mPAP, right ventricular systolic pressure (RVSP), mean right ventricular pressure (mRVP) and mean right atrial pressure (mRAP) were recorded. The blood counts, biochemical indexes, and coagulation parameters were collected for each patient. The parameters of blood count comprised of white blood cell (WBC), red blood cell (RBC), hemoglobin (HGB), platelets (PLT), variation of red blood cell distribution width (RDW-CV), standard deviation of red blood cell distribution width (RDW-SD), platelet distribution width (PDW), thrombocytocrit (PCT), mean platelet volume (MPV) and platelet-large cell rate(P-LCR). The biochemical indexes included creatinine (CRE), uric acid (UA), glutamic-pyruvic transaminase (ALT), glutamic oxalacetic transaminase (AST), gamma-glutamyl transpeptidase (GGT), C-reactive protein (CRP), N-terminal pro-brain natriuretic peptide (NT-proBNP) and troponin I (TNI). The coagulation test was routinely performed for all CHD patients by a standard clinical laboratory method in our hospital, the coagulation parameters including prothrombin time (PT), activated partial thromboplastin time (APTT), international normalized ratio (PT-INR), fibrinogen (FIB), thrombin time (TT), prothrombin time activity (PTA), antithrombin Ⅲ (AT-Ⅲ) and D-dimer (D-D) were extracted from medical records. Transthoracic echocardiography was routinely performed for all CHD patients and the echocardiographic parameters including left atrium end diastolic diameter (LA-Dd), left atrium end systolic diameter (LA-Ds), left ventricular end diastolic dimension (LV-Dd), right atrium end systolic diameter (RA-Ds), right ventricular end diastolic dimension (RV-Dd), main pulmonary artery (MPA), tricuspid regurgitation velocity (TAV) and tricuspid regurgitation pressure (TAP) were recorded. The other clinical data such as medical history and demographic information were also collected from the medical records.

### Blood+ -based proteomic analysis

The plasma from three groups: healthy people, patients with CHD and patients with CHD-PAH were used for the proteomic analysis. The plasma was processed by a series of steps, including protein extraction, trypsin digestion, LC-MS/MS analysis and bioinformatics analysis. Firstly, the cellular debris in the plasma were removed by centrifugation and high abundance proteins were removed by Pierce™ Top 14 Abundant Protein Depletion Spin Columns Kit (ThermoFisher Scientific). After determination of concentration by BCA kit, the extracted protein was digested by trypsin. Then, the digested proteins were desalted by C18 SPE column, followed by LC-MS/MS analysis. The resulting MS/MS data were processed by using MaxQuant search engine (v.1.6.15.0). Finally, the quantifiable proteins in coagulation cascade were extracted and the DEPs were identified by a threshold of p≤0.05.

### Enzyme-linked immunoassay analysis

Plasma proteins of coagulation cascade and markers of thrombin formation were quantified by enzyme-linked immunoassay (ELISA) kit (Cloud-Clone Corp., China) following the manufacturer’s instructions. Briefly, standards and samples are added to the microplate wells with a biotin-conjugated antibody. Next, avidin conjugated to horseradish peroxidase (HRP) is added to each microplate well and incubated. After TMB substrate solution is added. The enzyme-substrate reaction is terminated by the addition of sulphuric acid solution and the color change is measured by a microplate reader (Thermo Scientific, USA) at a wavelength of 450nm. The concentration of protein in the samples is then determined by comparing the O.D. of the samples to the standard curve. The evaluation of coagulation cascade was undertaken by measuring the protein levels of coagulation system, natural anticoagulant system and fibrinolytic system. The coagulation system was evaluated by measuring the protein levels of coagulation factors and their receptors, including Tissue Factor (TF), Coagulation Factor VII (F7), Coagulation Factor VIII (F8), Coagulation Factor IX (F9), Coagulation Factor X (F10), Coagulation Factor XI (F11), Coagulation Factor II (F2), von Willebrand Factor (vWF), Coagulation Factor V (F5), Coagulation Factor XII (F12), Coagulation Factor XIII A1 Polypeptide (F13A1), Fibrinogen Alpha Chain (FGA), Kininogen 1 (KNG1) and Protease Activated Receptor 4 (PAR4). The natural anticoagulant system was assessed by measuring the levels of Thrombomodulin (TM), Protein C (PROC), Protein C Receptor, Endothelial (PROCR), Protein S (PROS), Heparin Cofactor II (HCII), Antithrombin-Ⅲ (AT-Ⅲ) and Tissue Factor Pathway Inhibitor (TFPI). Fibrinolytic marker Plasminogen (Plg), Tissue Plasminogen Activator (tPA) and Plasminogen Activator, Urokinase (uPA) were measured to assess the activation of fibrinolysis, whereas Thrombin Activatable Fibrinolysis Inhibitor (TAFI), Plasminogen Activator Inhibitor 1 (PAI1), Alpha-2-Macroglobulin (a2M), Alpha-1-Antitrypsin (a1AT) and Alpha 2-Antiplasmin (a2PI) were measured to assess inhibition of fibrinolysis. Prothrombin fragments (F1+2), Thrombin/Antithrombin Complex (TAT) and Fibrinopeptide A (FPA) were measured as markers for thrombin formation.

### The quantitative PCR (qPCR) analysis

Total RNA was isolated from lung tissues using Trizol reagent (Life Technology, USA). The RNA concentration and purity were assessed using NanoDrop™ instruments (Thermo Scientific, USA). First-strand cDNA was synthesized by using the Transcriptor First Strand cDNA Synthesis Kit, according to the manufacturer’s protocol. The first-strand cDNA was used for qPCR. The quantification of mRNA expression was performed by using Light Cycler 96 (Roche, Switzerland). The relative quantification was performed by the comparative 2^−ΔΔCT^ method and expressed as fold changes. The forward and reverse primers for the amplification of each mRNA fragment were as follows: forward-5’-CTT CTT CGT ACA AGC CGT GAT T-3’, and reverse-5’-ACA GAG ATA TGG TCA GCA GGA T-3’ for TF; forward-5’-CCC AGA ATC ACG GAG AAC ATG-3’, and reverse-5’-ACC ACA CCT GTC AGA TAC CA-3’ for F7; forward-5’-GAT GGA GGT GCC CTA TGT GG-3’, and reverse-5’-TGC CGG TCA CAA AGT AGG TG-3’ for F10; forward-5’-TGC CAA GGA TGA CGA AGG TG-3’, and reverse-5’-GGC CAT ATC GGA CTG GTG TT-3’ for F13A1; forward-5’-TCA GTG TGT TGG GGA CGA TG-3’, and reverse-5’-AGA CGA GCC ACT TCA CAA GG-3’ for vWF; forward-5’-CCA GTC CAG AGC AAT TAG TGT ATT-3’, and reverse-5’-CGT GTC TTC AAG TCC TGT GTT-3’ for F8; forward-5’-CGG CGT TTG CTA TGA CCA AG-3’, and reverse-5’-CTG GTC GGA TGT CTT CTC GG-3’ for AT-*Ⅲ;* forward-5’-AAT TCA AGT ACG GCG GTT GC-3’, and reverse-5’-CGC TTT GGT AGC CTG AGG AAT-3’ for TFPI; forward-5’-GAG TTG GTG GAC GGA GAG TG-3’, and reverse-5’-GAA CAT TTC GCA CCT GTC GG-3’ for TM; forward-5’-TGG AGA ACC ACA TCA CCA CG-3’, and reverse-5’-GGC CAC ACC AGC GAT TAT GA-3’ for PROCR; forward-5’-GCA TCA TTG GAG ACA CGA GAG-3’, and reverse-5’-TGT AGA CGC CAT AGT TGT TGA G-3’ for PROC; forward-5’-CAG CTT GTC TGC CAT CTC CA-3’, and reverse-5’-TAG ATA CTC AGC GCG GTT GC-3’ for PLG; forward-5’-ATT CCA GTC AGT GTG CCC AG-3’, and reverse-5’-CCG TAG CCA GAA AGC TCA CA-3’ for tPA; forward-5’-AGC TGG GCA TGA CTG ACA TC-3’, and reverse-5’-TGC GGG CTG AGA CTA GAA TG-3’ for PAI1; forward-5’-CGT ACC ACC TCG TAA GCC AA-3’, and reverse-5’-CCA CCA AGT CCC AGA TCC AC-3’ for a2M; forward-5’-CAG CCA GTG AGT GAG CAA GCC-3’, and reverse-5’-ACA GCC TGC GAG TCT CCT CTG-3’ for a2PI; forward-5’-CTA CCA TGG GAA TGT GAG TGT C-3’, and reverse-5’-ATT CTC CTT GGA ACC TCT TGA G-3’ for F2; forward-5’-CAG AGA CAT CTT TGC CTC CTT A-3’, and reverse-5’-ATT CTT TGC CTT TCA CAT CGA C-3’ for F11; forward-5’-ATA TGG ATC GAC TGT GGA ATC C-3’, and reverse-5’-TCT TTT TCC ACG TGT AGT CGT A-3’ for TAFI; and forward-5’-CTG GAG AAA CCT GCC AAG TAT G-3’, and reverse-5’-GGT GGA AGA ATG GGA GTT GCT-3’ for GAPDH.

### Statistical analysis

Continuous variables were expressed as mean ± SD and categorical variables as number (percentage). Comparison of two groups was performed using unpaired t test or Chi-square test. Multiple comparisons to control were performed with one-way ANOVA and followed by the Tukey’s multiple comparisons test within groups. Chi-square test was used for categorical variables. GraphPad Prism 8 was used for statistical analysis. A value of p < 0.05 was regarded as statistical significance. Further details of the statistical analysis were provided in the figure legends.

## Results

### Study design and pathological findings in PAH models

We analyzed the global coagulation cascade in both animal models and PAH patients at transcriptomic and proteomic levels, the study design was showed in the Figure 1A. The pathological examination was performed in monocrotaline (MCT)- and Sugen-Hypoxia (SuHx)-induced PH models, respectively. On gross examination, the surface of lungs was diffusely congested with punctate hemorrhagic foci and hemorrhagic areas in MCT-induced PAH (Figure 1B). Histopathological examination of lung tissue showed pulmonary vascular remodeling, such as medial hypertrophy, laminar intimal proliferation and fibrosis with the consequence of lumen stenosis. Notably, one of the remarkable pathologic findings was thromboembolism and *in situ* thrombosis in the pulmonary vessels (Figure 1C). The thromboembolism and *in situ* thrombosis were frequently found in the hemorrhagic lung, in which fresh thrombi were identified. Similar to MCT-induced PAH, the hemorrhagic foci and areas were observed in the SuHx-induced PH, but with fewer abnormalities (Figure 1D). Microscopically, the thromboembolism and *in situ* thrombosis in the pulmonary vessel were also recognized in SuHx-induced PH (Figure 1E).

**Figure 1.**
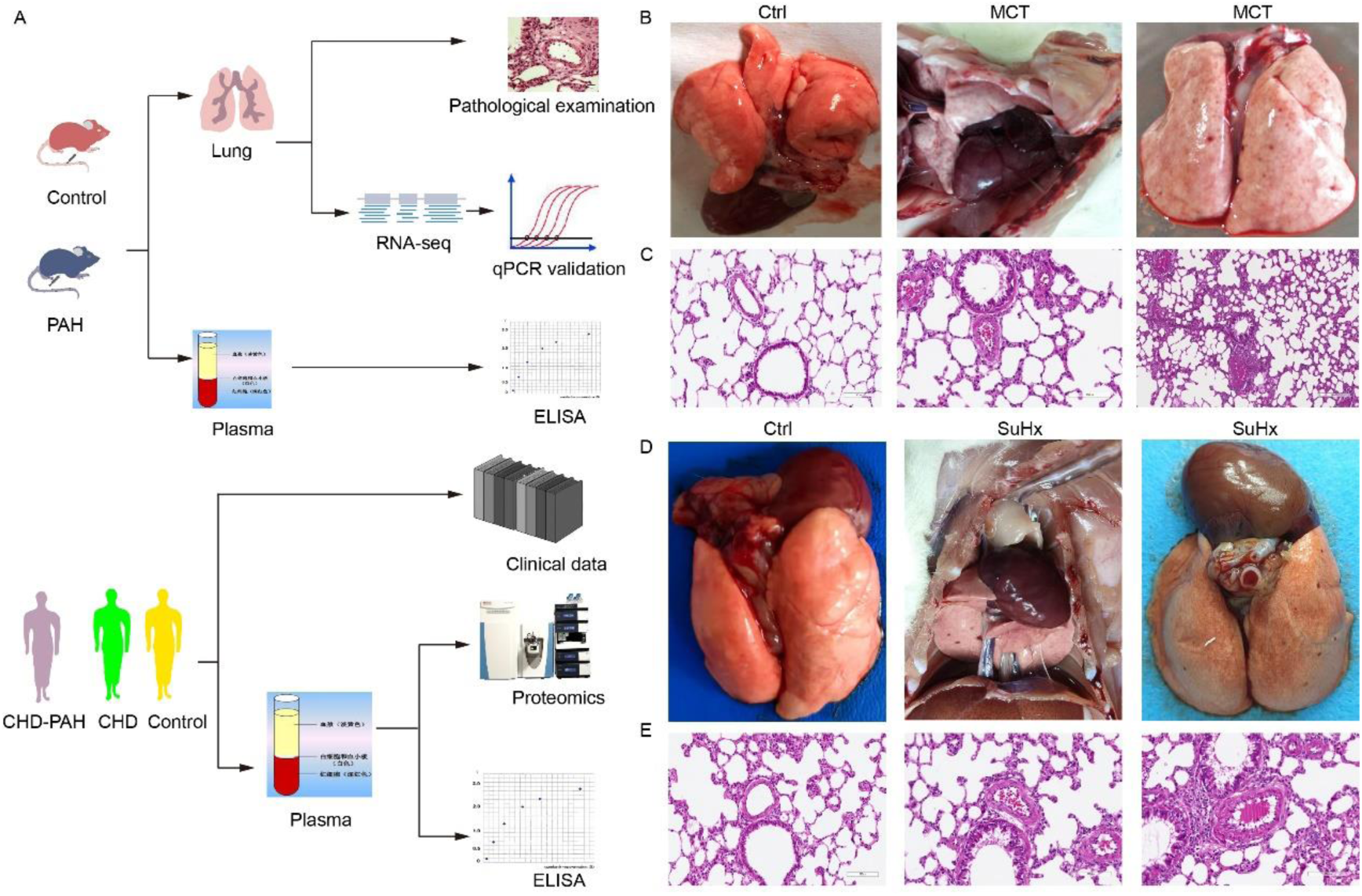
Study design and pathological examination of lung tissue. **A**, The experimental design of this study. Coagulation cascade in PAH animal model and patient was analyzed by using RNA sequencing, proteomic approach and ELISA. **B**, Gross morphology of control and MCT-treated lung. Pathological manifestations of hemorrhage on the lung surface. **C**, Representative images of lung hematoxylin and eosin (HE) staining in MCT-induced PAH. Pathological examination showed thromboembolism and *in situ* thrombosis. **D**, Gross morphology of control and SuHx-treated lung. Pathological manifestations of hemorrhage on the lung surface. **E**, Representative images of lung HE staining in SuHx-induced PH. Pathological examination showed thromboembolism and *in situ* thrombosis.

### Transcriptomic analysis of coagulation cascade in MCT-induced PAH

Our previous study has showed the sustained enrichment of complement and coagulation pathways in the progression of MCT-induced PAH ^23^. To explore the mechanisms of thrombosis and bleeding observed in the pathological examination, the genes annotated in coagulation cascade were extracted from transcriptomic datasets. In addition to F9 and Serpina5, all genes associated with coagulation cascade were identified by RNA sequencing (Data Set S1). Hierarchical clustering of coagulation system showed both reduced and increased expression patterns of the coagulation factors, and differentially expressed analysis showed the reduced expression of multiple intrinsic coagulation factors, including F11, F8 and Vwf, with the exception of increased common coagulation factor F10 (Figure S1A). In addition, transcriptional profiling of fibrinolytic system showed both fibrinolytic and anti-fibrinolytic genes were in an increased expression trend except a1AT (Figure S1B). Differentially expressed analysis revealed increased expression of fibrinolytic genes Plg, and tPA, whereas anti-fibrinolytic genes PAI1, a2M and TAFI were simultaneously upregulated, especially for the PAI1 whose elevation from the onset to progression of PAH (Figure S1B). Notably, transcriptional profiling of natural anticoagulant system showed reduced trend of almost all anticoagulants, differentially expressed analysis revealed downregulation of AT-Ⅲ, HCⅡ, TM, PROS and TFPI (Figure S1C), indicating impairment of three anticoagulant pathways. To confirm the transcriptomic data, we analyzed the coagulation cascade in an irreversible form of PAH model with GSE149899 dataset ^24^. We identified the consistent downregulation of F11, F8, TM and TFPI and upregulation of F10, tPA, Plg, TAFI and PAI1 in the irreversible PAH, moreover the expression of extrinsic and common clotting factor TF, F2, Fgb and Fgg were found to be significantly upregulated (Figure S1D). The qPCR was used to validate the differentially expressed genes (DEGs) identified by RNA sequencing. The qPCR analysis confirmed the reduced intrinsic factors F11 and vWF and increased F10, reduced anticoagulants TM and AT-Ⅲ, as well as increased PAI1, TAFI and a2M in MCT-induced PAH (Figure S1E).

### The change of plasma coagulation cascade in MCT-induced PAH

To determine whether coagulation cascade altered in the lung tissue was similarly altered in the peripheral circulation, the whole coagulation cascade in the plasma was assessed by ELISA. The coagulation system had a reduced trend except extrinsic TF, an initiator of extrinsic coagulation (Figure 2A and Figure S2A). Similar to increased mRNA levels in the lung, the plasma level of TF was elevated (Figure 2A). In contrast, we found significant reduction in both intrinsic and common clotting factors F5, F8, vWF, F9, F10, F11, F2 and KNG1 (Figure 2A), of which vWF, F8 and F11 were sustained reduction in the progression of PAH (Figure S2C). Thereby, these results revealed the impairment of both intrinsic and common pathways in the peripheral circulation. The natural anticoagulant systems had an increased trend with an exception of AT-*Ⅲ* and TFPI (Figure 2B and Figure S2A). Consistent with mRNA levels, the plasma levels of AT-*Ⅲ* and TFPI were significantly reduced in MCT-induced PAH (Figure 2B), and further ELISA assessment revealed a sustained reduction in plasma AT-*Ⅲ* activity from the initiation and progression of PAH (Figure S2C). In contrast to mRNA levels, the plasma levels of anticoagulant PROC, PROS and TM in protein C system were significantly elevated (Figure 2B). Moreover, the HC*Ⅱ*, a thrombin inhibitor, was also found to be elevated in the plasma (Figure 2B). The fibrinolytic activators had an increased trend, whereas fibrinolysis inhibitors had a reduced trend, except PAI1 (Figure 2C and Figure S2B). Likewise, ELISA analysis confirmed increased plasma levels of fibrinolytic activators uPA and Plg, as well as fibrinolysis inhibitor PAI1 in the plasma (Figure 2C). Moreover, the PAI1 level was sustained increase from the onset to progression of PAH (Figure S2C).

**Figure 2.**
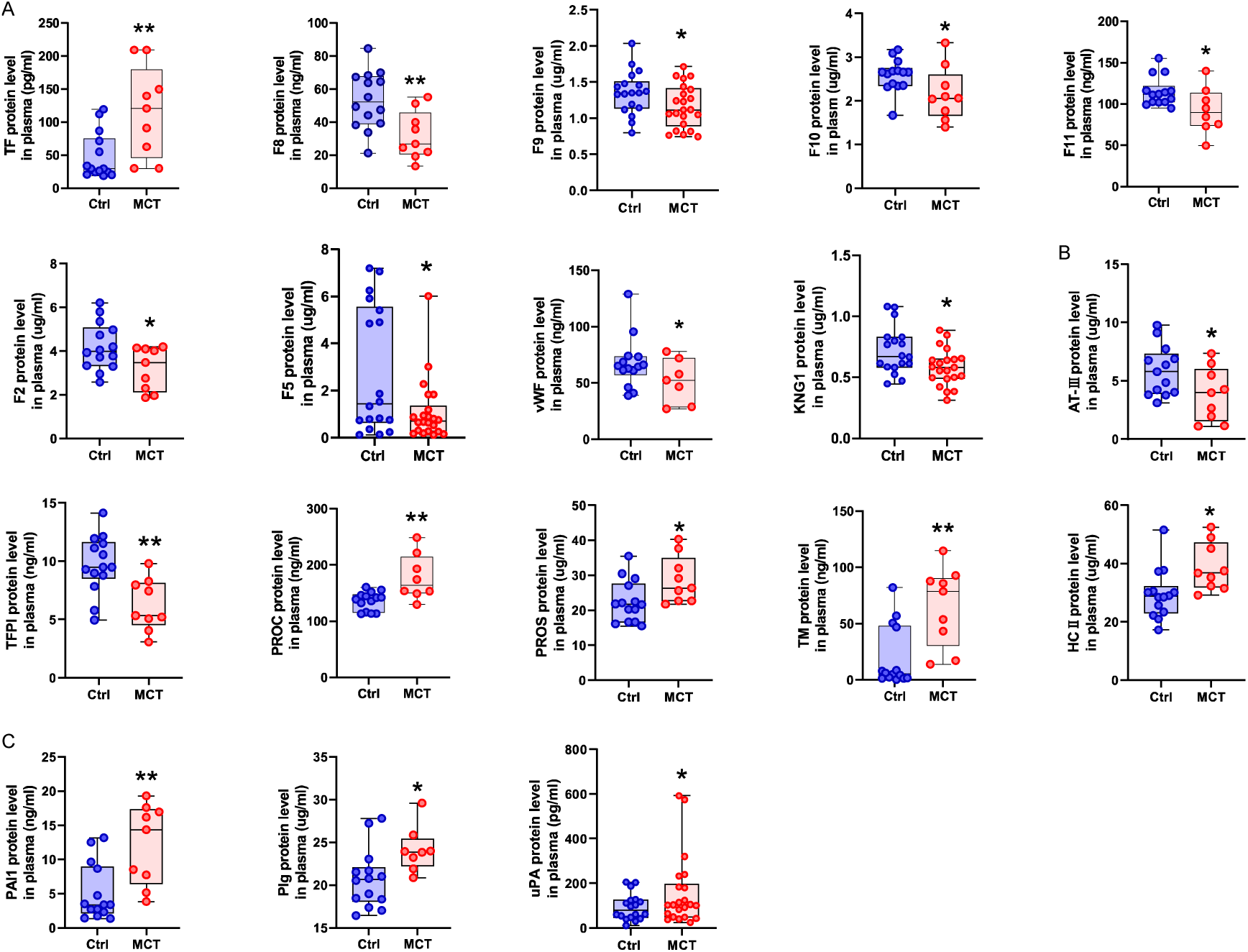
Plasma levels of coagulation cascade in MCT-induced PAH. **A**, The plasma levels of factors in coagulation system. **B**, The plasma levels of natural anticoagulants. **C**, The plasma levels of factors in fibrinolytic system. Box plot, centre line represents the median, box extends from the 25th to 75th percentile and whisker extend to the most extreme data points for consideration of outliers. Data were analyzed by unpaired t test (two-tailed), n=14 or 18 control, n= 9 or 22 MCT-treated rats at week 4. *p<0.05 vs control and **p<0.01 vs control. Ctrl, control; MCT, monocrotaline.

### Transcriptomic analysis of coagulation cascade in SuHx-induced PH

To investigate whether the altered coagulation cascade similarly occurred in the other PAH model, we performed RNA sequencing on lung tissues of SuHx-induced PH. A total of 25700 transcripts was identified, including 23255 in control and 24989 in SuHx group, and PCA analysis showed the SuHx samples were not completely separated from control samples (Figure 3A). Moreover, only 471 genes were differentially expressed in SuHx group, including 353 upregulated and 118 downregulated genes (Figure 3A), which were less than the numbers identified in MCT-induced PAH using the same threshold ^23^. All genes annotated in coagulation cascade were identified except for F9, F13b and Serpina5 (Data Set S2). Hierarchical clustering analysis of coagulation cascade generated two clusters of expression pattern (Figure 3B). One cluster with a trend to reduce was mainly inclusive of anticoagulants and intrinsic coagulation factors. The other cluster with an increased expression trend was composed of factors in fibrinolytic system, and coagulation factors mainly in extrinsic and common coagulation pathways. Despite a nearly complete coagulation cascade was identified in the lung, only a few numbers of DEGs were identified. As showed in the Figure 3C, differentially expressed analysis showed the reduced intrinsic clotting factors F8 and F11 and increased F10. Additionally, we identified downregulated TM, a1AT and PAR4, as well as upregulated a2M, TAFI and PAI1 (Figure 3C). To confirm the altered coagulation cascade in the SuHx-induced PH, we analyzed the coagulation cascade using an independent dataset, GSE8078. Differential expression analysis showed reduced intrinsic factors F8, F9 and vWF and increased extrinsic TF and KNG1 in hypoxia- and SuHx-induced PH (Figure 3D). Consistently, we identified the downregulated TFPI and simultaneously upregulated tPA and PAI1 (Figure 3D). Likewise, validation of DEGs by qPCR confirmed the reduced expression of vWF, TFPI, TM and PROC, and increased TF, F10, PAI1 and a2M (Figure 3E). Together, similarly altered coagulation cascade occurred in the SuHx-induced PH model, but showed fewer abnormalities.

**Figure 3.**
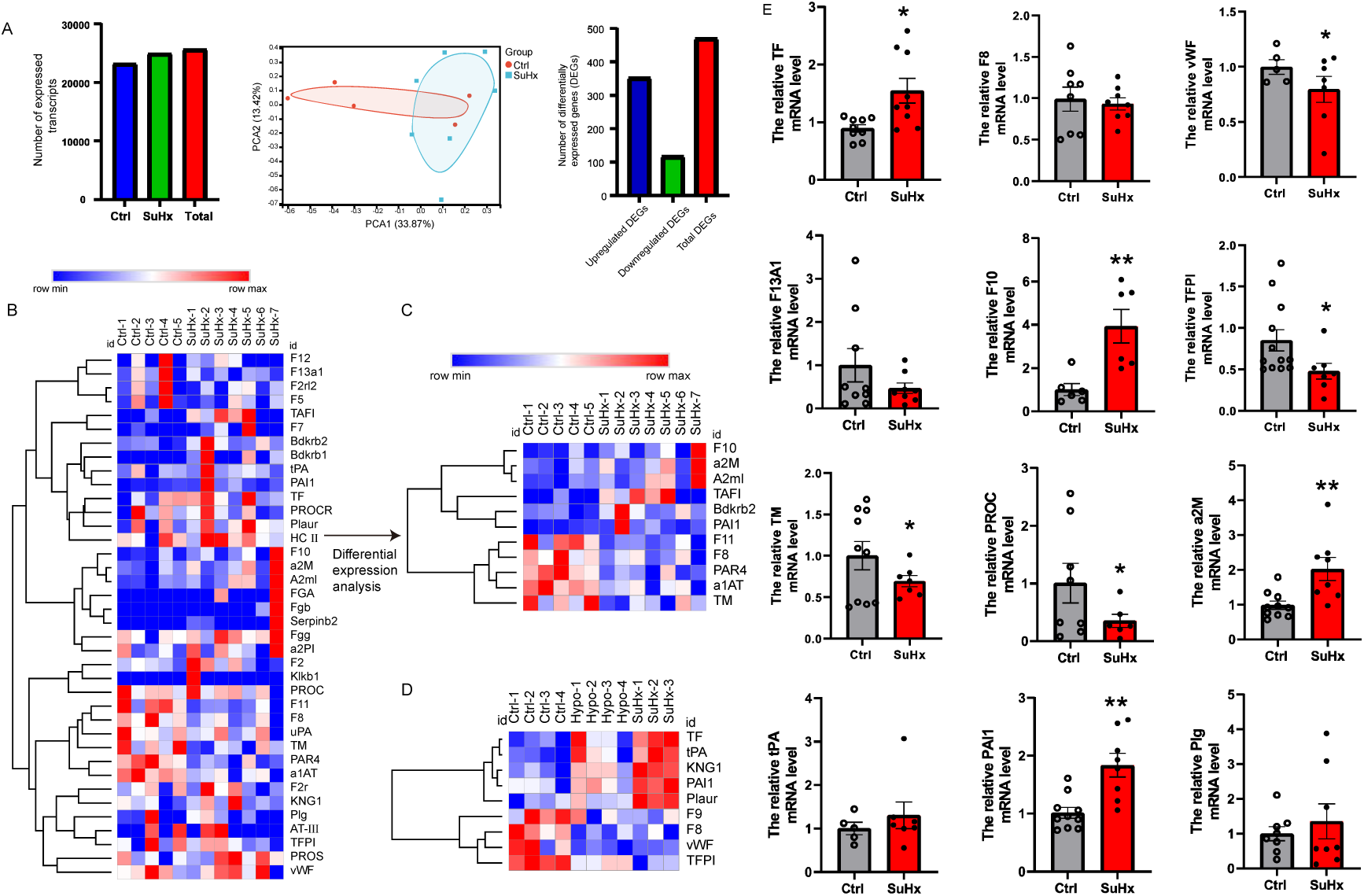
Transcriptional profiling of coagulation cascade in lung of SuHx-induced PH. **A**, The transcriptome and DEGs identified by RNA sequencing in SuHx-induced PH. **B**, Hierarchical clustering analysis of the coagulation cascade. **C,D**, Heatmap of DEGs in the coagulation cascade. Rows in the heatmap represent gene expression levels and columns represent each sample. The DEGs between control and SuHx group were identified by using a threshold of p≤0.05. **E**, qPCR validation of the DEGs expression in coagulation cascade. Data were expressed as mean ± SD and analyzed by unpaired t test (two-tailed), n=6-12 control, n= 6-9 SuHx. *p<0.05 vs control and **p<0.01 vs control. Ctrl, control; SuHx, Su5416+Hypoxia.

### The change of plasma coagulation cascade in the SuHx-induced PH

ELISA assessment of coagulation system in the peripheral circulation showed that several coagulation factors had a reduced trend in the SuHx-induced PH (Figure 4A and Figure S3A). Compared with control, the plasma levels of intrinsic and common clotting factors F8, F11, FGA and KNG1 were significantly reduced, whereas TF, vWF and PAR4, a thrombin receptor for platelet activation, were elevated (Figure 4A). The natural anticoagulant systems in the plasma had an increased trend except AT-*Ⅲ* and TFPI (Figure 4B and Figure S3B). In comparison with control, the plasma levels of AT-*Ⅲ* and TFPI were significantly reduced in SuHx-induced PH (Figure 4B). Similar to protein C system in MCT-induced PAH, the plasma levels of anticoagulants TM, PROC and PROS were elevated (Figure 4B), which was not in accordance with mRNA expression in the lung tissue. In addition, the fibrinolytic system had a reduced trend except PAI1, tPA and uPA (Figure 4C and Figure S3C). As compared with control, we found the reduced plasma level of TAFI and increased PAI1 in the fibrinolytic system (Figure 4C).

**Figure 4.**
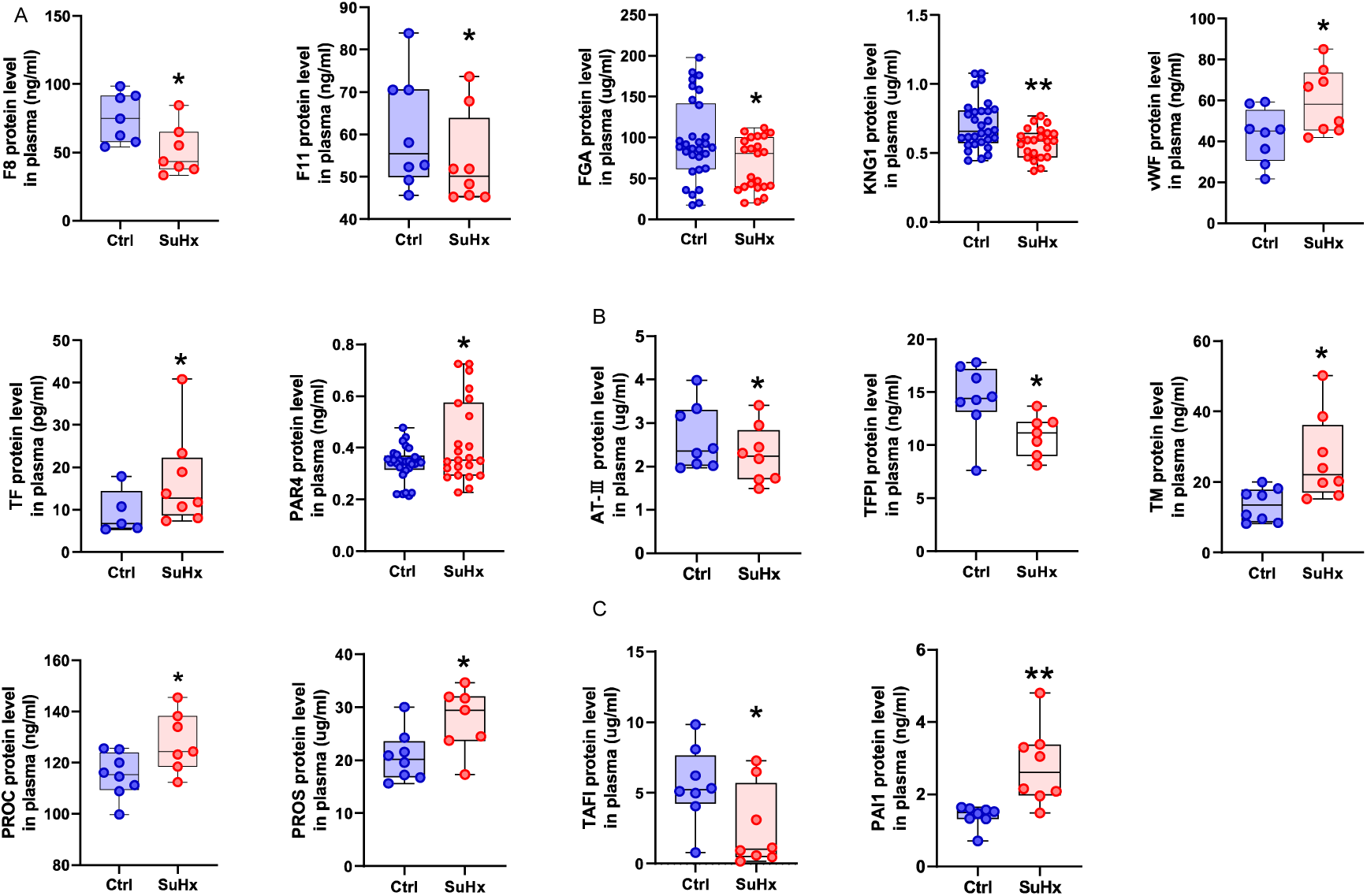
Plasma coagulation cascade in SuHx-induced PH. **A**, The plasma levels of factors in coagulation system. **B**, The plasma levels of natural anticoagulants. **C**, The plasma levels of factors in fibrinolytic system. Box plot, centre line represents the median, box extends from the 25th to 75th percentile and whisker extend to the most extreme data points for consideration of outliers. Data were analyzed by unpaired t test (two-tailed), n=8 or 30 control, n= 8 or 23 SuHx. *p<0.05 vs control and **p<0.01 vs control. Ctrl, control; SuHx, Su5416+Hypoxia.

### Hemostatic abnormalities in patients with CHD-PAH

The baseline characteristics for the subjects were summarized in Table 1. There were no statistical significances in age, gender, WBC and RBC, *etc.* among three groups. In comparison with control subjects, the values of RDW-CV, AST, GGT, NT-proBNP and CRP were elevated, while HGB, PLT and PCT were reduced in patients with CHD-PAH. In addition to elevated hemodynamic parameters mPAP, RVSP and mRVP, CHD-PAH patients showed elevated echocardiographic measurements such as RA-Ds, RV-Dd and MPA, elevated biochemical variables AST, GGT and NT-proBNP, as well as abnormal blood count including reduced PLT, PCT and increased RDW-CV as compared with CHD patients (Table 1).

**Table 1.**
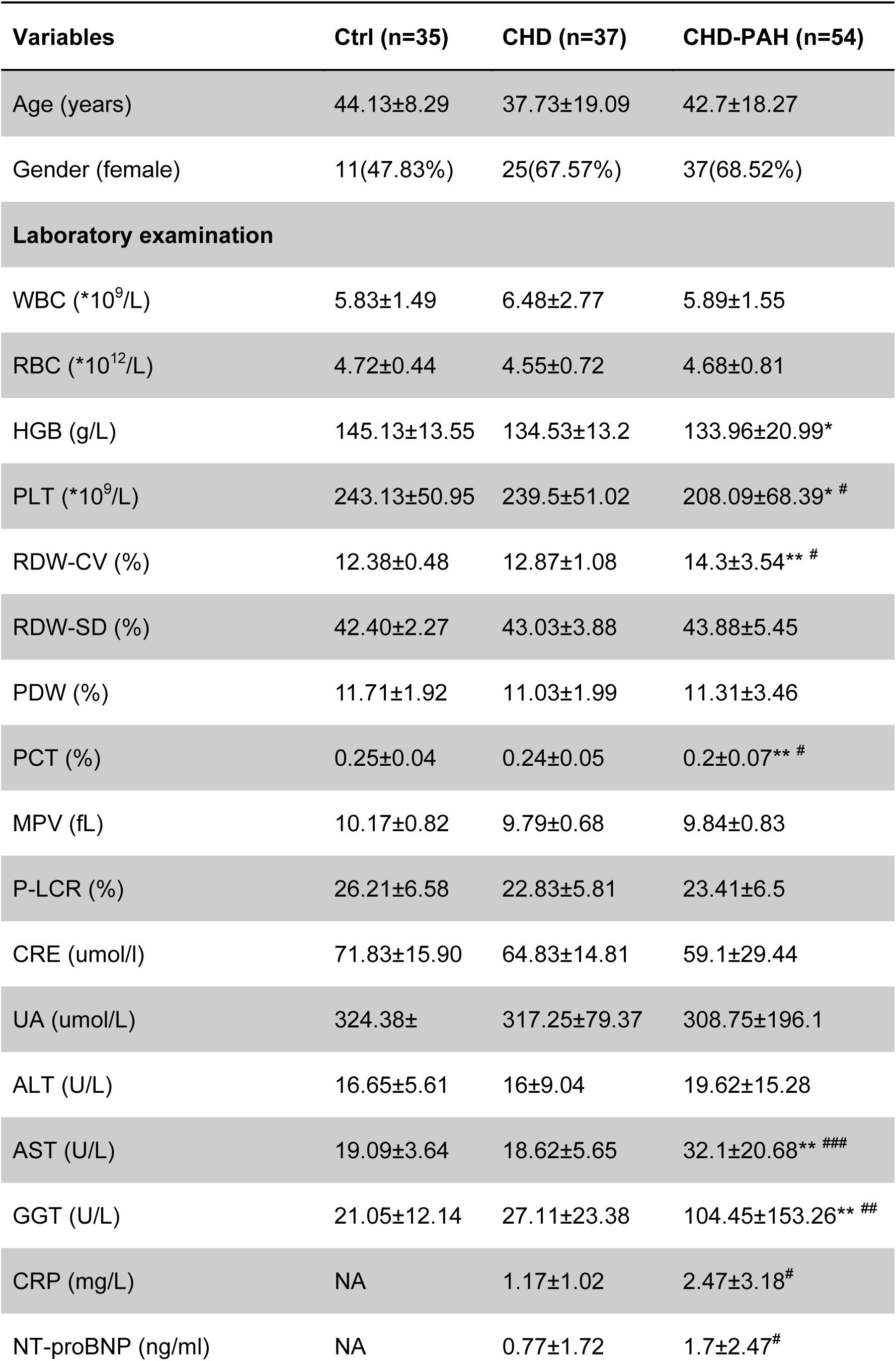
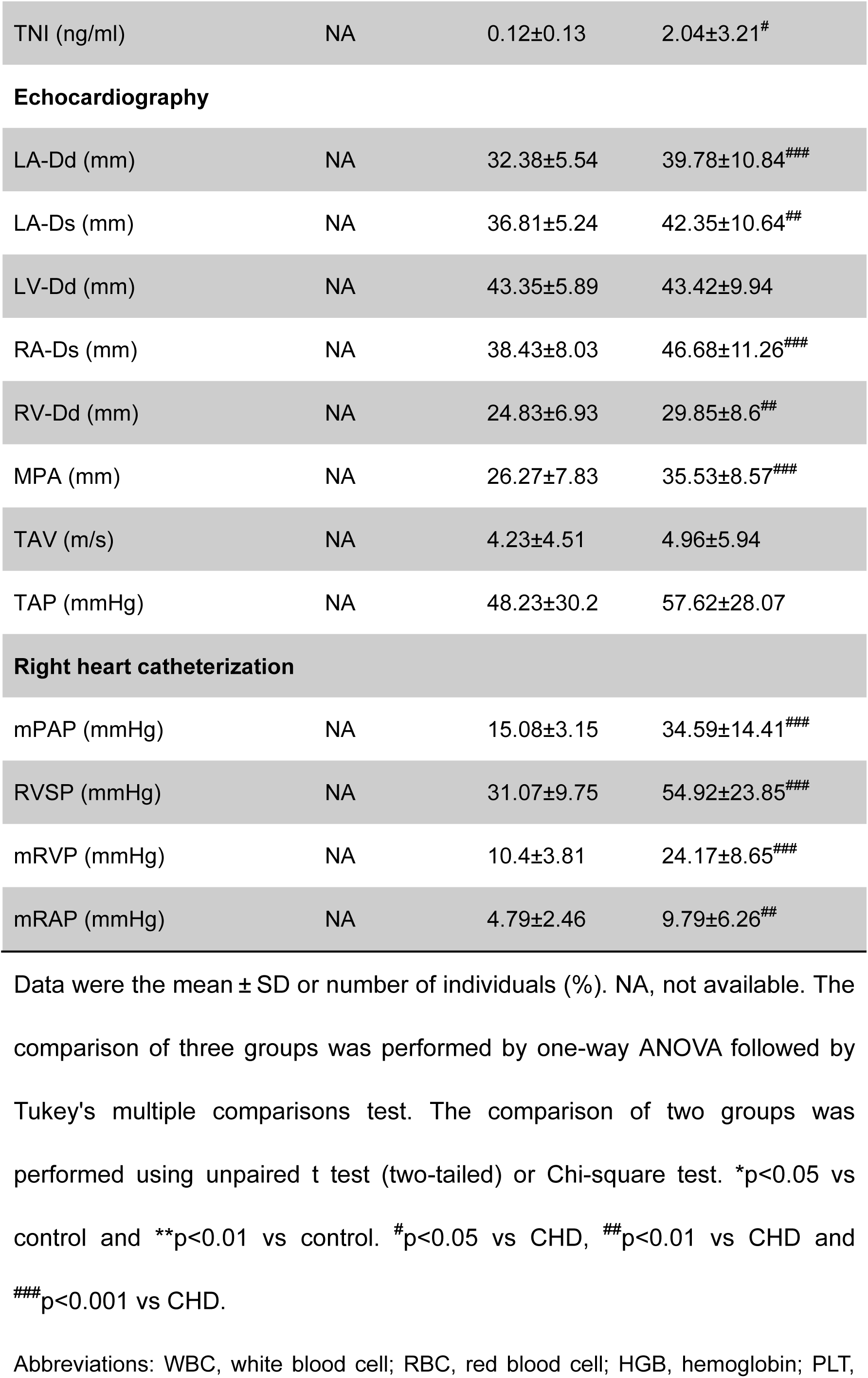
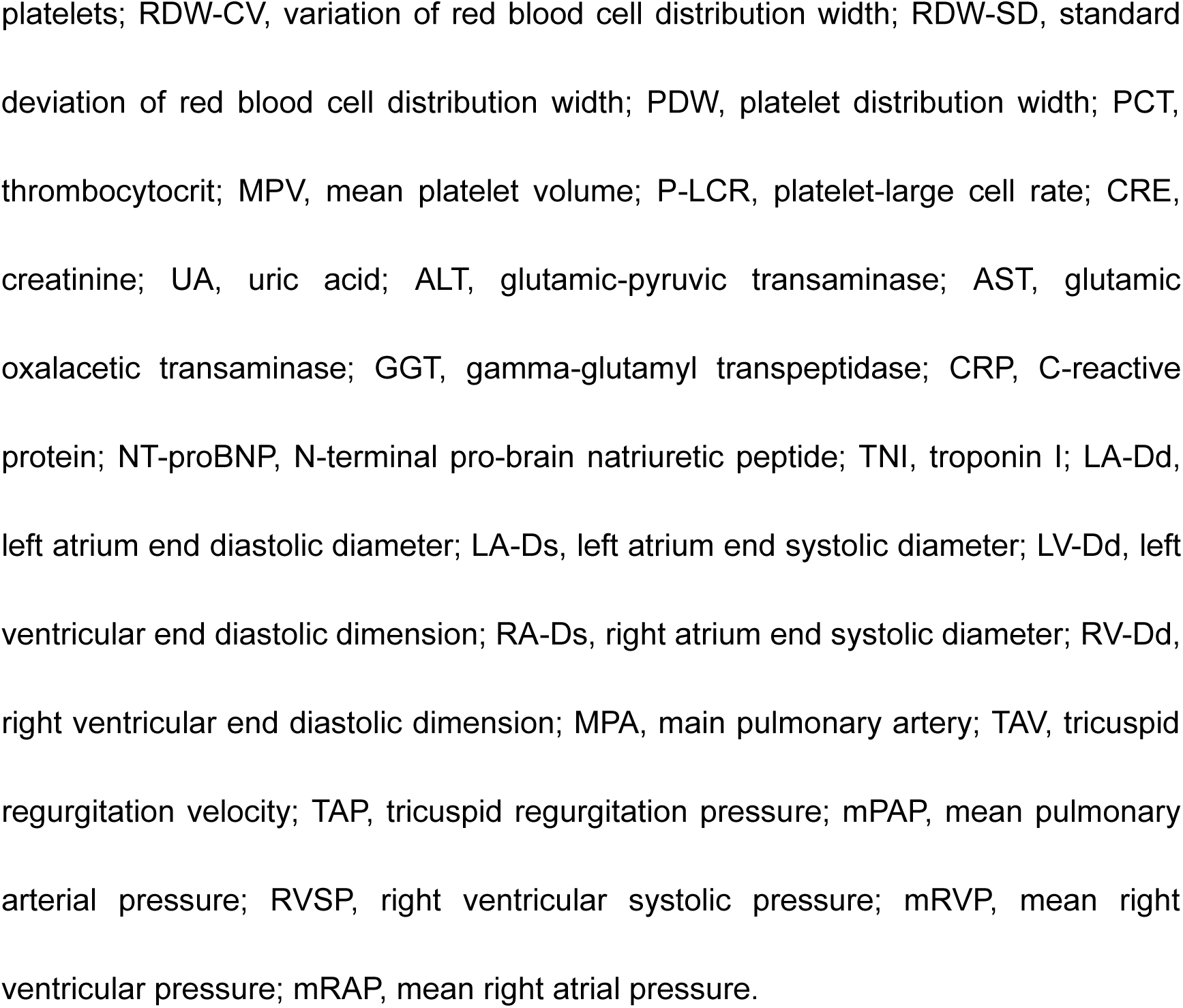
Baseline clinical characteristics for the subjects.

In comparison with control subjects, CHD-PAH patients presented higher values of PT, INR and TT in spite of increased D-dimer (Figure 5), reflecting the lower activity of coagulation factors. Additionally, the activities of PTA and AT-Ⅲ were reduced in CHD-PAH patients. As a result, the prolonged, but not shortened, hemostatic variables PT, TT and INR and reduced PTA indicated the hypocoagulability and hemostatic deficiency in CHD-PAH patients. Likewise, prolonged values of coagulation variables PT, INR and APTT were found in CHD-PAH patients as compared with CHD patients, whereas activities of PTA and AT-Ⅲ were reduced (Figure 5). Of note, the majority of AT-Ⅲ activity were in a low reference range in patients with CHD-PAH (Figure 5). Moreover, compared with PAH patients with normal AT-Ⅲ activity, PAH patients with lower AT-Ⅲ activity had prolonged PT and INR, decreased PTA, PCT and PLT, as well as increased NT-proBNP, CRE, UA and ALT (Table S1). Consequently, these results indicated the lower AT-Ⅲ activity was associated with defective hemostasis and exacerbated cardiac, renal and liver function.

**Figure 5.**
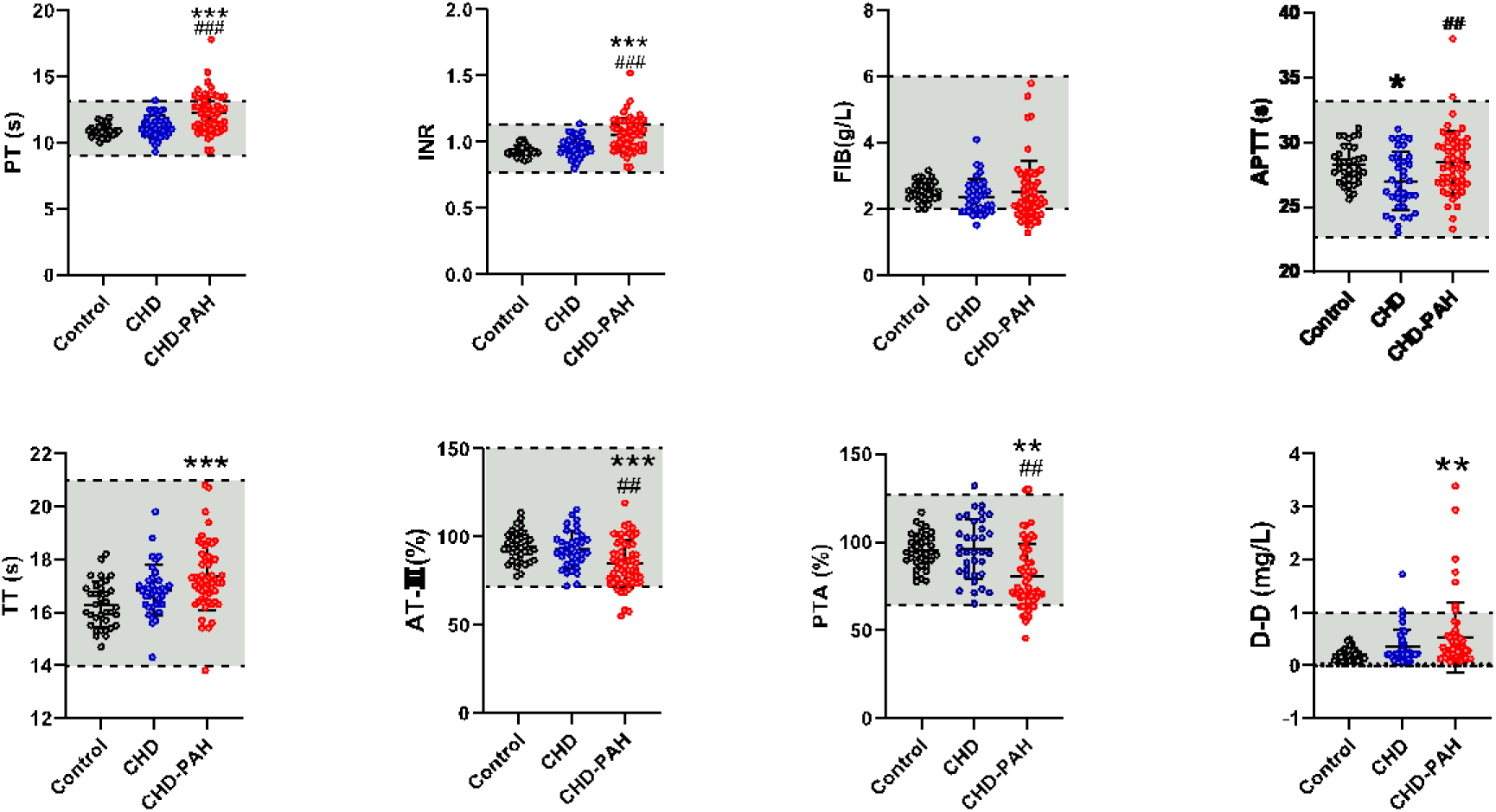
Hemostatic abnormalities in patients with CHD-PAH. Individual and mean values of coagulation parameters were shown in the figure. Shaded area denotes normal reference range. Data were showed as mean ± SD and analyzed by one-way ANOVA followed by Tukey’s multiple comparisons test. n=35 control, n= 37 CHD, and n= 54 CHD-PAH. *p<0.05 vs control, **p<0.01 vs control and ***p<0.001 vs control. ^#^p<0.05 vs CHD, ^##^p<0.01 vs CHD and ^###^p<0.001 vs CHD. CHD, congenital heart disease; CHD-PAH, congenital heart disease-associated PAH. PT, prothrombin time; APTT, activated partial thromboplastin time; PT-INR, international normalized ratio; FIB, fibrinogen; TT, thrombin time; PTA, prothrombin time activity; AT-Ⅲ, antithrombin Ⅲ; D-D, D-dimer.

### Proteomic analysis of coagulation cascade in patients with CHD-PAH

To identify the plasma protein profiles of coagulation cascade in patients with CHD-PAH, we used blood+ -based proteomic approach to obtain a view of coagulation cascade. A total of 1493 proteins were identified, of which 1294 were quantifiable (Figure 6A). PCA analysis of proteome showed that CHD-PAH patients were separated from CHD and control subjects (Figure 6B). Blood+ -based proteomic analysis showed that 25 proteins of coagulation cascade were quantifiable, accounting for 65.85% proteins and the majority of proteins have a trend to be reduced (Figure 6C). Proteomic profiling of coagulation cascade in patients with CHD-PAH showed reduced trend in plasma levels of anticoagulants, clotting factors and factors in fibrinolytic system with the exception of F7, a1AT and a2M, *etc.* (Figure 6D). As showed in the Figure 6E, 10 of 25 proteins were differentially expressed in CHD-PAH group as compared with control. The reduced plasma proteins were intrinsic and common clotting factors F11, F10, F2 and KNG1, anticoagulants AT-Ⅲ and PROS, as well as a2PI and Plg in fibrinolytic system. Notably, the plasma levels of extrinsic F7 and a1AT were increased in CHD-PAH group (Figure 6E). In comparison with control, only 6 of 25 proteins were differentially expressed, including reduced F13A1, F13B, AT-Ⅲ and a2PI, and increased F7 and a1AT in CHD group (Figure 6F).

**Figure 6.**
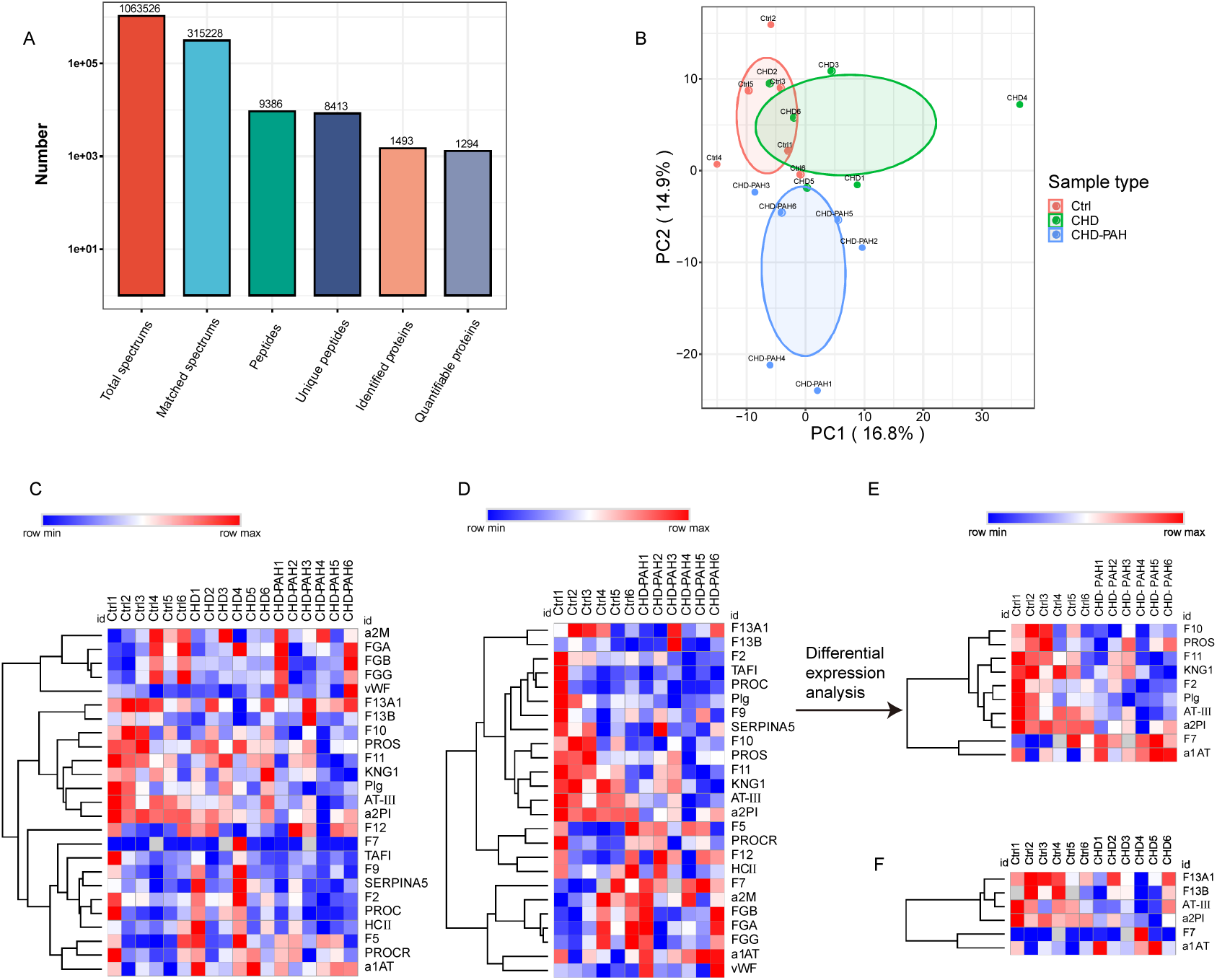
Proteomic analysis of coagulation cascade in patients with CHD-PAH. **A**, The quantifiable proteins by blood+ -based proteomic analysis. **B**, PCA analysis of proteome. **C,D**, Hierarchical clustering of coagulation cascade. **E,F**, Heatmap of differentially expressed proteins in coagulation cascade. Rows in the heatmap represent gene expression level and columns represent each sample (n=6). The differentially expressed proteins between control and CHD or CHD-PAH group were identified by using a threshold of p≤0.05. Ctrl, control; CHD, congenital heart disease; CHD-PAH, congenital heart disease-associated PAH.

### The abnormalities of coagulation cascade in patients with CHD-PAH

Further ELISA assessment of coagulation system showed both the increased and reduced trend of coagulation factors (Figure 7A and Figure S4A). As compared with control subjects, plasma levels of extrinsic TF and F7, and intrinsic F8 were significantly higher in patients with CHD-PAH (Figure 7A). With the exception of TF, F7 and F8, the other clotting factors, including intrinsic and common F10, F11, F2 and vWF were significantly reduced in CHD-PAH patients compared with control subjects (Figure 7A), suggesting impairment of both intrinsic and common pathways in the peripheral circulation. Similar to PAH models, the anticoagulant systems had an increased trend with the exception of AT-Ⅲ, PROS and HCII (Figure 7B and Figure S4B). Compared with control, the plasma levels of AT-*Ⅲ* and PROS were significantly reduced in patients with CHD-PAH (Figure 7B). In contrast to AT-Ⅲ level, the anticoagulant TFPI was increased in the peripheral circulation (Figure 7B). There was a trend to increase in the protein C anticoagulant system except PROS. However, only PROCR, an endothelial protein C receptor, was found to be elevated (Figure 7B and Figure S4B). The fibrinolytic inhibitors showed a reduced trend except for PAI1 and a1AT (Figure 7C and Figure S4C). In comparison with controls, fibrinolysis inhibitors a2PI and TAFI were significantly lower in CHD-PAH patients (Figure 7C). By contrast, the fibrinolytic activators tPA and uPA showed an increased trend (Figure 7C and Figure S4C). Despite both plasma levels of PAI1 and tPA were significantly elevated in patients with CHD-PAH, the value of PAI1/tPA was not elevated (Figure 7C). Consequently, these results did not support fibrinolytic inhibition in patients with CHD-PAH. Notably, CHD patients had increased levels of F7 and reduced F10, F2, F5, AT-Ⅲ, PROS, TAFI and a2M as compared with control subjects, while CHD-PAH patients had increased levels of TF, TFPI, tPA and FGA, as well as reduced vWF, TAFI and a2PI in comparison with CHD patients, indicating CHD patients complicated with PAH had aggravated coagulation abnormalities (Figure 7 and Figure S4).

**Figure 7.**
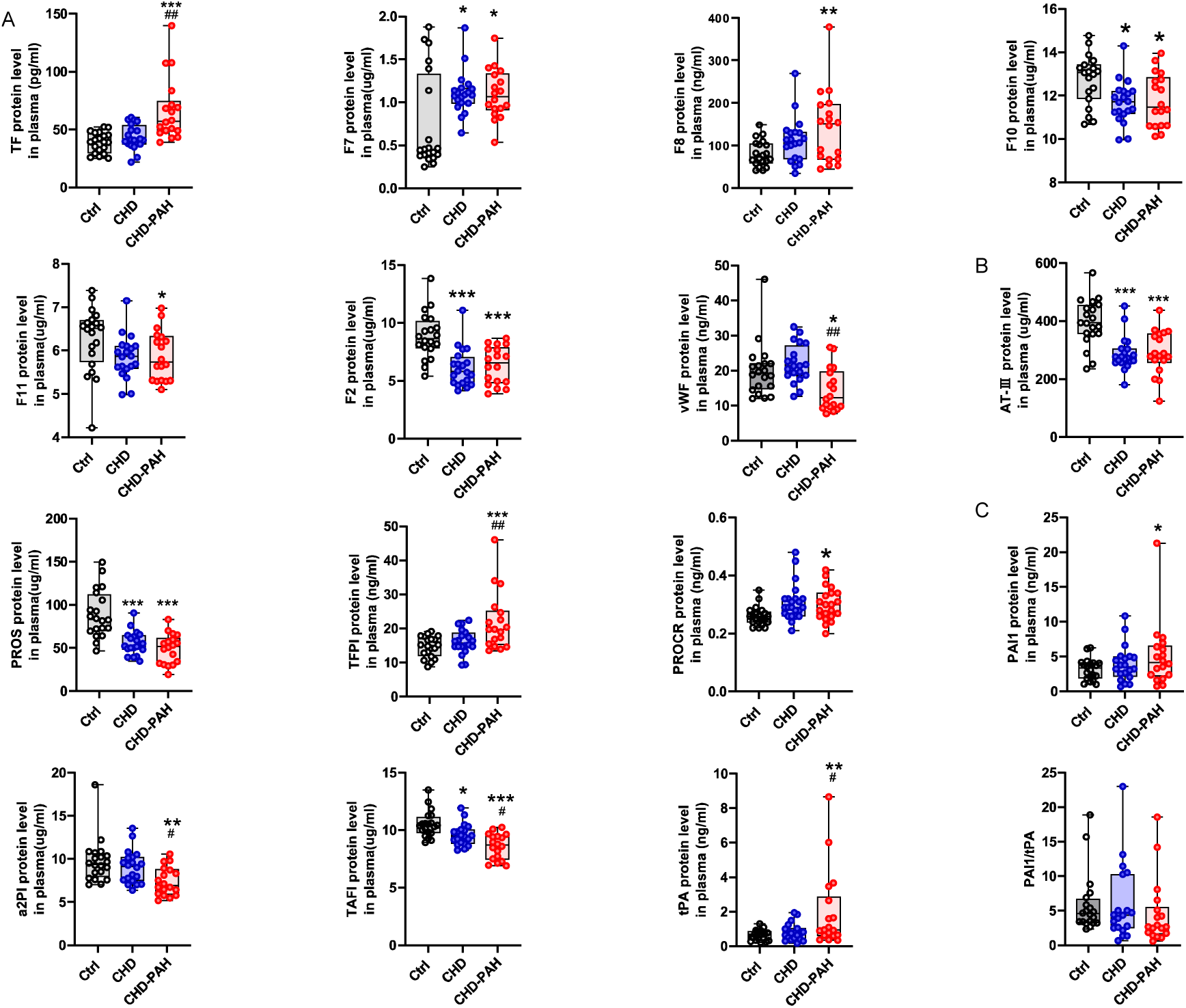
Plasma coagulation cascade in patient with CHD-PAH. **A**, The plasma levels of clotting factors. **B**, The plasma levels of factors in anticoagulation system. **C**, The plasma levels of factors in fibrinolytic system. Box plot, centre line represents the median, box extends from the 25th to 75th percentile and whisker extend to the most extreme data points for consideration of outliers. Data were analyzed by one-way ANOVA followed by Tukey’s multiple comparisons test, n= 21 control, n= 23 CHD, n= 21 CHD-PAH. *p<0.05 vs control, **p<0.01 vs control and ***p<0.001 vs control. ^#^p<0.05 vs CHD and ^##^p<0.01 vs CHD. Ctrl, control; CHD, congenital heart disease; CHD-PAH, congenital heart disease-associated PAH.

### The abnormal markers of thrombin formation in PAH

Patients with PAH are at a high risk for thromboembolic complications. To determine the thrombin generation in PAH, we assessed the plasma markers of thrombin formation by ELISA. In spite of a trend to increase in plasma levels of TAT and FPA in SuHx-induced PH, no statistically significant change was found (Figure S5A). The plasma levels of TAT and F1+2 have a trend to be elevated in MCT-induced PAH. However, FPA level was significantly reduced (Figure S5B). Notably, F1+2 and TAT were significantly decreased in patients with CHD-PAH and CHD as compared with controls and there was no significant change in the FPA level Figure S5C). Thus, as a whole, these results did not support increased thrombin formation in the peripheral circulation.

## Discussion

The majority of studies focus on individual abnormal factors of coagulation cascade in PAH. However, *in situ* thrombosis of pulmonary vessels and thromboembolic complications were involved in a series of components and their sequential interactions in the coagulation cascade. In the present study, we assessed the global coagulation cascade change in both lung tissues and peripheral circulation. We identified a nearly complete profiling of coagulation cascade at both the transcriptomic and proteomic levels, which may provide sights into the mechanisms underlying *in situ* thrombosis of pulmonary vessel and thromboembolic complication, respectively.

The PAI1 mRNA expression in the lung tissue was increased from the onset to progression of PAH. In addition to PAI1, the other antifibrinolytic genes TAFI and a2M were also elevated in the lung tissue. The increased antifibrinolytic genes may result in the impaired ability to remove the thrombus. However, the fibrinolytic genes such as tPA and Plg, were simultaneously elevated in PAH lung. Therefore, the overall fibrinolytic homeostasis appeared to be in a balance in the PAH lung. The fibrinolytic system comprised of eight factors and only a few numbers of factors were assessed in the previous studies. Both fibrinolysis inhibitor PAI1 and fibrinolytic activator tPA were reported to elevate in the plasma, accounting for the abnormality of fibrinolysis in patient with PAH^9, 10^. The evaluation of whole fibrinolytic system confirmed the elevated plasma levels of PAI1 and tPA in the present study. Notably, PAI1/tPA was not elevated in CHD-PAH patients and the other fibrinolysis inhibitors, such as TAFI and a2PI were reduced in the plasma. Consequently, these results do not support a fibrinolytic deficit and it was likely that presence of a functional fibrinolytic and antifibrinolytic balance in the peripheral circulation.

The extrinsic pathway is triggered by exposure TF to blood. TF interacts with F7 to form a TF-F7a complex and then activates common pathway, whereas TFPI is the main inhibitor of the TF-F7a complex. The increased TF expression has been reported in plexiform lesions from PAH rat models and patients^21^. In this study, we confirmed the increased TF expression and moreover, we identified the reduced expression of TFPI and increased common clotting factors F10 and F2 in PAH lung. Thereby, *in situ* thrombosis of the pulmonary vessels may be associated with activation of extrinsic pathway. Conversely, we observed diffuse hemorrhagic spots on the lung surface in pathological examination. The intrinsic pathway comprised of F12, F11 and F8, *etc.* plays an important role in maintaining and amplifying coagulation function. In contrast to extrinsic pathway, we identified the reduced expression of multiple intrinsic coagulation factors, such as F11, F8 and vWF in the PAH lung, thus indicating that impaired intrinsic pathway contributed to hemorrhage on the lung surface.

There are three anticoagulant systems: protein C system, TFPI system and heparin/AT-Ⅲ system. The protein C system is composed of PROC, PROS, TM and PROCR. The expression of TM, PROC and PROS were reduced in the PAH lung, thus suggesting the impairment of protein C anticoagulant system for *in situ* thrombosis of pulmonary vessels. The heparin/AT-Ⅲ system comprised of heparin and AT-Ⅲ was the predominant anticoagulant systems involved in the negative regulation of coagulation activation. In the present study, a reduction in AT-Ⅲ mRNA expression was identified in MCT-induced PAH, indicating an impairment of heparin/AT-Ⅲ anticoagulant system for *in situ* thrombosis of pulmonary vessels. To our knowledge, this is the first report to show impairment of heparin/AT-Ⅲ anticoagulant system in the PAH lung. CHD patients were at a high risk for thromboembolic complications including deep venous thromboembolism, pulmonary thromboembolism and stroke^5, 25^, and thromboembolism resulted in 8% cause-specific mortality in patients with Eisenmenger syndrome, an advanced form of CHD-PAH^6^. Paradoxically, the hemostatic deficits were frequently reported in CHD-PAH patients. The incidence of hemoptysis was reported as 3.1% to 5.5% in patients with CHD-PAH, and markedly increased in patients with Eisenmenger syndrome^17^. However, little was known about the mechanisms underlying thrombotic and/or bleeding complications.

Abnormalities in plasma clotting factors have not been adequately characterized in PAH patients except for vWF, F8 and fibrinogen with conflicting results^14, 22, 26^. All clotting factors were evaluated in the present study. We found an increase in the plasma levels of extrinsic TF and F7 in CHD-PAH patients. In contrast, the plasma levels of intrinsic and common clotting factors F10, F11, F2 and vWF were reduced. As a result, the thromboembolic complications in CHD-PAH were associated with activation of extrinsic pathway, but not intrinsic pathway. Studies assessing markers of thrombin formation in PAH have rendered inconsistent results with some studies showing increased and the others decreased thrombin formation ^8, 12, 13^.

We did not find the increased markers of thrombin formation. Instead, the markers of thrombin formation TAT, F1+2 were reduced in patients with CHD-PAH. The reduction in multiple coagulation factors in intrinsic and common pathways may provide an explanation for hemostatic deficiency and hemorrhagic events in patients with CHD-PAH. Despite the hemostatic deficit was recently explained by ongoing consumption ^12, 13^, the identification of raised clotting factors TF, F7 and F8 did not support this notion.

The protein C anticoagulant system exerts its anticoagulant function by inactivating clotting factors F5a and F8a in the presence of cofactor PROS. Contrary to mRNA expression in lung tissue, the plasma levels of anticoagulants TM, PROC and PROS were elevated in PAH rat models, reflecting activated protein C system. Of note, the activated protein C system may enhance anticoagulation and facilitate bleeding in peripheral circulation. In despite of a trend to increase in the protein C system in CHD-PAH patients, only PROCR was elevated and PROS was found to be reduced in the peripheral circulation. Consequently, the abnormalities in protein C system may not play a substantial role in thrombotic complications. The conflict results in TFPI level were reported previously, because both increased and reduced TFPI levels have been demonstrated in patients with PAH ^8, 27^. Similarly inconsistent results were showed in the present study, both reduced plasma levels of TFPI in animal models and increased TFPI in CHD-PAH patients were found. In contrast to protein C and TFPI systems, the AT-Ⅲ plasma level was consistently reduced in both MCT-induced and SuHx-induced PH and furthermore, AT-Ⅲ plasma level was also reduced in CHD-PAH patients, indicating that impairment of heparin/AT-Ⅲ anticoagulant system was an evolutionarily conserved mechanism for thrombosis across rat and human PAH.

The transcriptional profile of coagulation cascade altered in lung tissue was largely consistent with corresponding coagulation cascade altered in the peripheral circulation, thus the abnormalities of coagulation cascade in plasma may be mainly originated from diseased lung tissue. However, it was not consistent in terms of protein C system and common pathway. A limitation of this study is that the exact mechanisms underlying these inconsistent results remained unclear, we cannot exclude a correlation with impaired liver function due to elevated biochemical variables AST, GGT in CHD-PAH patients. The reduction in synthesis of intrinsic clotting factors and anticoagulants, such as AT-Ⅲ and TFPI, was observed in the PAH lung. The second limitation is that the regulatory mechanisms remained unknown. The reduced synthesis of intrinsic coagulation factors in the lung may contribute to low-procoagulant state and haemostatic defects, whereas the impairment of anticoagulant system in the lung may result in a low-anticoagulant state and increased tendency to develop *in situ* thrombosis. As a result, we proposed that a new functional balance of lower procoagulant and anticoagulant factors may exist to reduce the thrombosis in the pulmonary circulation. However, this new balance is vulnerable and easily distorted by a variety of factors, including warfarin, new oral anticoagulants, inflammation and infection, thereby predisposing PAH patients to thrombosis, bleeding, or even both.

In summary, impairment of heparin/AT-Ⅲ anticoagulant system and activation of extrinsic pathway were conserved mechanisms for thrombosis in both experimental and human PAH. Since the heparin/ AT-Ⅲ anticoagulant system was severely impaired, restoring its function with heparin supplementation may be a better option for future anticoagulant therapy, but currently there was no clinical trial evaluating the safety and efficacy of heparin in PAH treatment.

## Acknowledgments

We thank all the investigators and participants in this study.

## Supplemental Material

Figure S1-S5

Tables S1

Data Set S1-S2

